# Clinical Validation of Optical Genome Mapping for the Detection of Structural Variations in Hematological Malignancies

**DOI:** 10.1101/2022.12.27.22283973

**Authors:** Andy Wing Chun Pang, Karena Kosco, Nikhil Sahajpal, Arthi Sridhar, Jen Hauenstein, Benjamin Clifford, Joey Estabrook, Alex Chitsazan, Trilochan Sahoo, Anwar Iqbal, Ravindra Kolhe, Gordana Raca, Alex R. Hastie, Alka Chaubey

## Abstract

Structural variations (SVs) play a key role in the pathogenicity of hematological malignancies. Standard-of-care (SOC) methods such as karyotyping and fluorescence *in situ* hybridization (FISH), employed globally for the past three decades have significant limitations in the resolution or the number of recurrent aberrations that can be simultaneously assessed, respectively. Next-generation sequencing (NGS) based technologies are now widely used to detect clinically significant sequence variants but are limited in their ability to accurately detect SVs. Optical genome mapping (OGM) is an emerging technology enabling the genome-wide detection of all classes of SVs at a significantly higher resolution than karyotyping and FISH. OGM neither requires cultured cells nor amplification of DNA and hence addresses the limitations of culture and amplification biases. This study reports the clinical validation of OGM as a laboratory developed test (LDT), according to CLIA guidelines, for genome-wide SV detection in different hematological malignancies. In total, 68 cases with hematological malignancies (of various subtypes), 27 controls and two cancer cell lines were used for this study. Ultra-high molecular weight DNA was extracted from the samples, fluorescently labeled, and run on the Bionano Genomics Saphyr system. A total of 207 datasets, including replicates, were generated and 100% could be analyzed successfully. Sample data were then analyzed using either disease specific or pan-cancer specific BED files to prioritize calls that are known to be diagnostically or prognostically relevant. Accuracy, precision, PPV and NPV were all 100% against standard of care results. Sensitivity, specificity, and reproducibility were 100%, 100% and 96%, respectively. Following the validation, 11 cases were run and analyzed using OGM at three additional sites. OGM found more clinically relevant SVs compared to SOC testing due to its ability to detect all classes of SVs at much higher resolution. The results of this validation study demonstrate OGM’s superiority over traditional SOC methods for the detection of SVs for the accurate diagnosis of various hematological malignancies.

## Introduction

Hematological malignancies refer to a distinct group of neoplastic diseases of hematopoietic and lymphoid tissues, and are broadly divided into myeloproliferative neoplasms, myelodysplastic neoplasms, leukemias, lymphomas, and plasma cell neoplasms. Historically, these malignancies have been genetically characterized for diagnosis, classification, prognostication, and therapeutic decision-making^1-3^. In the recent decade, the genetic testing of these malignancies has dramatically evolved with the advancement of genomic technologies, particularly molecular characterization due to the advent of next-generation sequencing (NGS) technology^4-6^. Though there have been significant improvements in molecular characterization of these blood tumors with NGS, the evolution of cytogenetic analysis has lagged comparatively, which still relies on traditional methods including karyotyping, Fluorescence in-situ hybridization (FISH), and chromosomal microarray (CMA).

Karyotyping (KT), currently the “gold standard” cytogenetic method for the detection of single-cell genome-wide structural variation (SV), suffers from several limitations: 1) limited resolution of aberration sizes (10-20 Mbp), 2) cryptic translocations that remain undetected, and 3) lack of metaphase cells in certain malignancies that result in failed KT (e.g., CD138+ cells in plasma cell myeloma, lymph node single cell suspension in lymphoma)^7,8^. FISH is a targeted assay often performed alongside KT or in isolation (for CD138+, lymph node single cell suspension or disease monitoring) to capture a limited number of SVs. CMA has seen limited adoption for hematological malignancies, despite it being ideal for copy number variation (CNV) detection, since it fails to detect insertions, balanced SVs (translocations, and inversions) or fusions^9^. The aforementioned limitations of these cytogenetic/cytogenomic methodologies necessitate the use of multiple assays to obtain a reasonable cytogenetic profile in majority of these cases^3,10,11^. Recently, NGS has been explored as a method to detect cytogenetic aberrations^12^, but the intrinsic limitation of poor sequencing around and within repetitive sequences of the genome result in limited resolution and inability to detect SVs^13^. NGS performs very well for the detection of sequence variants of clinical relevance but requires multiplexing, complex bioinformatics, high depth of coverage, and high costs associated with its implementation for the detection of cytogenetic aberrations. However, NGS is the perfect tool to complement OGM for a comprehensive genomic analysis^17^.

Recently, multiple studies have demonstrated that optical genome mapping (OGM) is a promising next-generation cytogenomic technology that provides a streamlined workflow at a high precision for the detection of all classes of SVs at a genome-wide level. These studies have shown ∼100% concordance of OGM with classical cytogenetic methods (KT, FISH and CMA) in specific hematological malignancies^14-18^. In addition, OGM has demonstrated the ability to detect additional clinically relevant SVs missed by SOC owing to its significantly higher sensitivity and resolution (∼10,000× compared to KT).

In this multi-site, IRB-approved OGM study on hematological malignancies, the clinical validation was conducted for the development of a laboratory developed test (LDT) in CLIA certified laboratories. The samples were evaluated for concordance, reproducibility, and assay robustness and protocols were established for the analysis and interpretation using guidelines-based targeted variant assessment in addition to a whole genome analysis. The unique ability of OGM to detect all classes of balanced and unbalanced SV at high resolution and increased sensitivity holds promise for its acceptance as a first tier cytogenomic test for all hematological malignancies. This is in line with multiple published studies demonstrating the reproducibility and robustness of the OGM workflow as it simplifies the clinical testing laboratory operations.

## Methods

### Cohort design

Multiple U.S.-based laboratories contributed to this double-blinded observational study for sample recruitment, data collection, and variant analysis. The study protocol was approved through multiple Institutional Review Boards (IRBs) and included consent provided by individuals with newly collected samples or waived authorization for use of de-identified samples. All protected health information (PHI) was removed, and data were anonymized (coded and double-blinded) before accessioning for the study. Samples were given anonymous aliases used in this study (e.g.: BNGOHM-xxxxxxx). All clinical samples (peripheral blood or bone marrow aspirate, N=71) were referred for clinical cytogenetic testing due to a suspicion of a hematological malignancy and SOC test results including karyotyping, FISH, and/or CMA were available. Control specimens included either established cell lines, peripheral blood, and bone marrow samples from healthy adults. For the de-identified cases, clinical indications, genetic test results and additional demographic information, as available, were collected.

### OGM Assay Workflow (DNA isolation, DNA Labeling, Chip loading and Data collection)

Frozen aliquots of cells, bone marrow aspirate (BMA), and/or peripheral blood (PB) were subjected to DNA isolation using manufacturer’s protocol (Bionano Genomics, Inc, USA). Briefly, frozen sample vials were thawed in a 37°C water bath, then counted for number of cells using the HemoCue WBC Analyzer (Fisher Scientific). A total of 1.5 million cells per sample was transferred to Protein Lo-Bind microfuge tubes for centrifugation. Cell pellets were resuspended and washed with stabilizing buffer. Washed cell suspensions were enzymatically digested and lysed, and isopropanol used to precipitate ultra-high molecular weight (UHMW) DNA in the presence of a nanobind disk. Long DNA strands bound to nanodisks were washed, transferred to clean tubes, and subsequently released from the nanodisk using elution buffer.

Approximately 500ng-750ng of solubilized UHMW DNA was labeled enzymatically, conjugating fluorophores to the target 6-mer *CTTAAG*. Long, labeled DNA strands were then counterstained with an intercalating dye, homogenized in buffer to allow flow through a nanochannel device, and then loaded into flowcells of Saphyr G2.3 chips. Chips were run in the Saphyr instrument to a target throughput of >1500 Gbp per sample.

### Assay QC

The completed datasets were then assessed for the following analytical quality control metrics — >320X effective coverage of GRCh38, with ≥70% of molecules ≥150 kbp aligning (“map rate”) and at an N50 of ≥230 kbp. Additionally, the Bionano Access 1.7 EnFocus™ (Fragile X) pipeline was run for a subset of samples to assess post-analytical QC pass/fail metrics (CNV noise and stable region analysis) and to infer sex of the case.

### SV detection using rare variant pipeline

The Bionano Solve 3.7 rare variant pipeline was used for genome-wide SV detection. The rare variant pipeline enables the detection of SVs occurring at low allelic fractions. Molecules were aligned to the GRCh38 reference, and clusters of molecules (≥3) indicating SVs were used for local assembly. Local consensus assemblies have high accuracy and are used to make final SV calls by realignment to the reference genome. SV calls were finally compared against known genes and against SVs in an SV database with 179 population controls.

Separately, based on read depth, the analysis generates a copy number profile that can call gains and losses in an approach analogous to microarrays. Briefly, molecules were aligned to the GRCh38 reference to create a depth-of-coverage profile, which was then normalized based on OGM controls and scaled against a baseline defined at CN=2 in autosomes (X and Y have a sex chromosome-specific baseline). Putative copy changes were segmented, and calls were generated and similarly annotated with positional information from the original reference. Entire chromosomal aneusomies were likewise defined in the CN algorithm.

### Post-Analytical SV Curation and Classification

A SV filtering and curation protocol was devised and implemented by analysts using Bionano Access v1.7.2. An overview of the subsequent curation and classification procedure is shown in Supplementary Figure 1. For the concordance part of the study, a set of filters was applied to include variants with a variant allele fraction (VAF) 0.02-1 and present in 0% controls of OGM control sample SV database. SVs meeting these criteria were then curated for manual review and classification. The analyst remained blinded until the concordance was performed. SVs were classified using a tiering system based on the ACMG guidelines adopted for OGM. Curation and classification were performed in four successively applied-then-removed filtering steps: disease subtype-specific classification (first, when applicable; most stringent filter), pan-hematological malignancy classification, pan-cancer classification, and remaining variants classification (last and most permissive filter) (Supplementary Figure 1).

Following validation, cases were run and analyzed according to SOP in an end-to-end exercise. In these cases, after variant analysis and preliminary classification, draft reports were reviewed by board certified pathologists or laboratory directors for final classification, interpretation, and readiness for future reporting. Directors proceeded through classified variants in the curated list, upholding and/or revising analyst classification, as needed. Upon completing review of the curated/classified variant list, the directors finalized case summary statements regarding somatic variants and defined genome complexity status (defined as normal if no large (≥5 Mbp) aberrations were detected, simple if fewer than 3 were detected, and complex for cases containing ≥3 aberrations).

### Concordance with SOC

Concordance analysis was performed between SVs detected using SOC testing and OGM data. Most of the samples had KT or KT plus FISH SOC results, and a subset had CMA data. If the same SV was observed in more than one SOC method, they were considered a single SV for concordance purposes. A SV was scored as concordant if the chromosome and band matched. Any difference in size of breakpoints were attributed to technique differences and the higher resolution of OGM. Additionally, if multiple OGM calls supported one SOC variant, they were treated as one concordant event. In addition to the concordance assessment, each case was evaluated for additional pathogenic/likely pathogenic (tier 1 and 2) findings that were not detected by SOC. Orthogonal methods were used to confirm a subset of these additional SVs detected by OGM.

### Reproducibility

UHMW DNA from four hematological malignancy BMA samples was labeled in different batches by different operators using multiple reagent lots and instruments. Reproducibility for each individual SV was calculated as the number of replicates where the SV was detected divided by the total number of replicates. Reproducibility for each SV type was calculated by taking the mean value across all variants for each type.

### Limit of detection

The limit of detection (LOD) was determined using two acute myelogenous leukemia (AML) cell lines, KG-1 and MV4-11 (ATCC). Each cell line DNA was blended with GM12878 (normal control cell line, Coriell Institute) to create serial dilutions that ranged from undiluted down to 1:24 dilution. Combined samples were gently mixed over the course of one week to ensure uniform mixing of DNA molecules. Six replicates of each blend were run through OGM with a target of >1500 Gbp of DNA and analyzed with the rare variant pipeline.

## Results

In this study, samples from 68 cases with hematological malignancies (with various heme subtypes), two cancer cell lines, and 27 controls were used (n=97, Table 1). From these clinical samples, cell lines, and controls, 207 datapoints were generated. Molecule N50, map rate, and effective coverage were recorded as analytical quality metrics, and they averaged 261 kbp, 87%, and 434×, respectively (Supplementary Figure 2).

**Table 1.** Summary of samples used for validation studies. * Phenotypically healthy individuals

| Sample type | Collection tube | Count | Normal control or clinical sample | Sample description |
| --- | --- | --- | --- | --- |
| BMA | Na-Heparin | 3 | Normal* | Frozen specimen taken through the entire wet lab process from extraction to OGM |
|  |  | 9 | Clinical sample |  |
|  | EDTA | 54 | Clinical sample |  |
| Blood | EDTA | 24 | Normal* |  |
|  | Na-Heparin | 2 | Clinical sample |  |
| UHMW gDNA | N/A | 3 | Clinical sample | UHMW gDNA |
| ATCC cell line | N/A | 2 | Cell line from AML cases | UHMW gDNA |
| **TOTAL** |  | **97** |  |  |

**Table 2.** Summary of concordance results. 2A) Concordance of hematological variants by sample type. 2B) Concordance by variant class.

| Sample Type | Total Samples Analyzed | Total Abnormal Events | Total Abnormal Concordant | Concordance |
| --- | --- | --- | --- | --- |
| BMA | 39 | 134 | 134 | 100% |
| WB | 2 | 3 | 3 | 100% |
| Cell lines | 2 | 11 | 11 | 100% |

| SV type by SOC testing | OGM SV |  |  | Percent Agreement |
| --- | --- | --- | --- | --- |
|  | TP | FN | Total |  |
| Aneuploidies | 61 | 0 | 61 | 100% |
| Gains | 8 | 0 | 8 | 100% |
| Deletions/losses | 51 | 0 | 51 | 100% |
| Translocations | 20 | 0 | 20 | 100% |
| Inversion | 1 | 0 | 1 | 100% |
| Add | 4 | 0 | 4 | 100% |
| Isochromosome | 1 | 0 | 1 | 100% |
| Dicentric | 1 | 0 | 1 | 100% |
| Idic | 1 | 0 | 1 | 100% |
| **Total** | **148** | **0** | **148** | **100%** |

Concordance with SOC testing results was conducted using 41 hematological malignancy cases (39 BMA and 2 blood, Figure 1), two hematologic malignancy cell lines and 27 normal healthy donor samples (3 BMA and 24 blood samples). Altogether, 148 SV calls by SOC testing were evaluated, and OGM demonstrated 100% concordance (Figures 2A and 2B). In addition, in 17/46 cases (37%) OGM identified additional novel Tier 1 and 2 variants not previously reported by SOC; a subset of these variants was subsequently confirmed by orthogonal methods (Supplementary Table 2). In one case of a myeloid neoplasm, BNGOHM-0000149, OGM was able to uniquely identify a mosaic deletion of 3q13.31 to 3q22.3 overlapping *GATA2*, a gene associated with myelogenous leukemia and an inclusion criterion for at least one clinical trial (NCT01861106), and complex rearrangements on 17p13, consisting of amplifications, and deletions that impacted *YWHAE, MNT, TP53* and *MAP2K4* (Figure 2A). In a second case of acute lymphocytic or myeloid leukemia, BNGOHM-0000335, OGM uniquely detected a translocation between chromosomes 16 and 12 [t(12;16)(p13;p13)] which results in the *CREBBP-ZNF384* fusion gene, an abnormality reported recurrently in ALL (Figure 2B).

**Figure 1.**
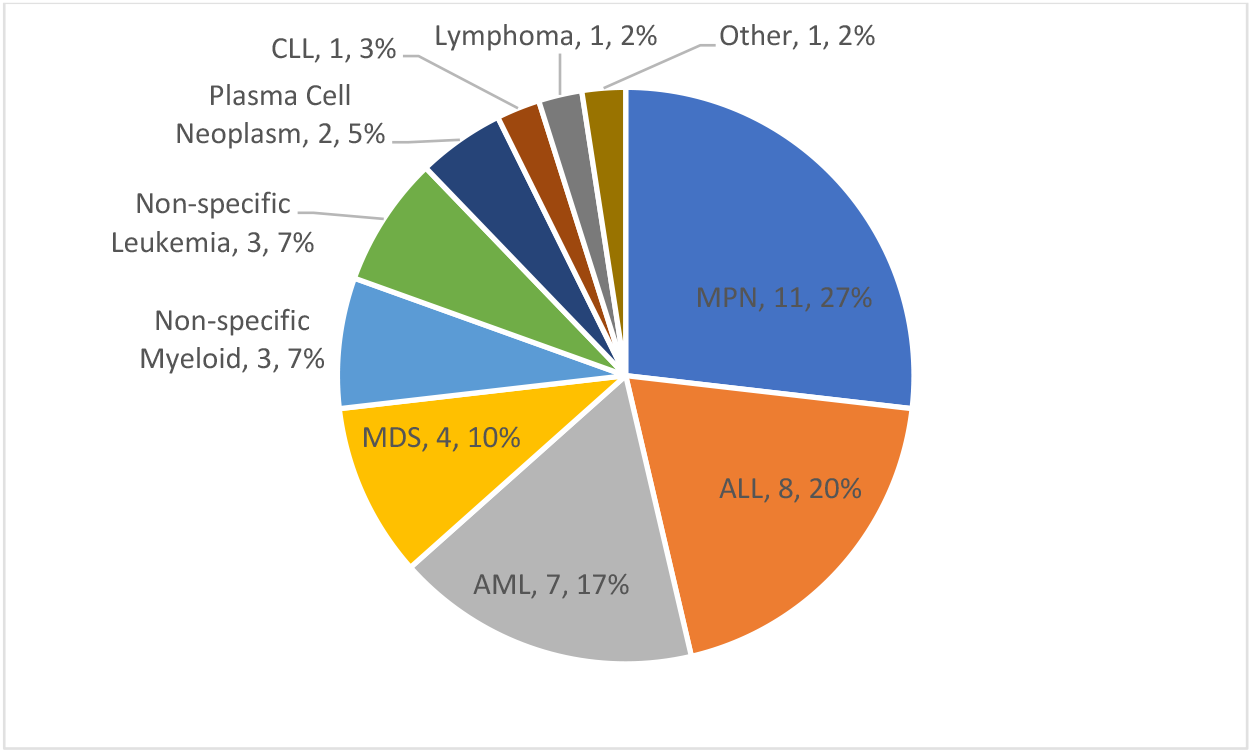
A breakdown of clinical indications of the cases used in validation for accuracy assessment (n=41, 2 cell lines not included).

**Figure 2.**
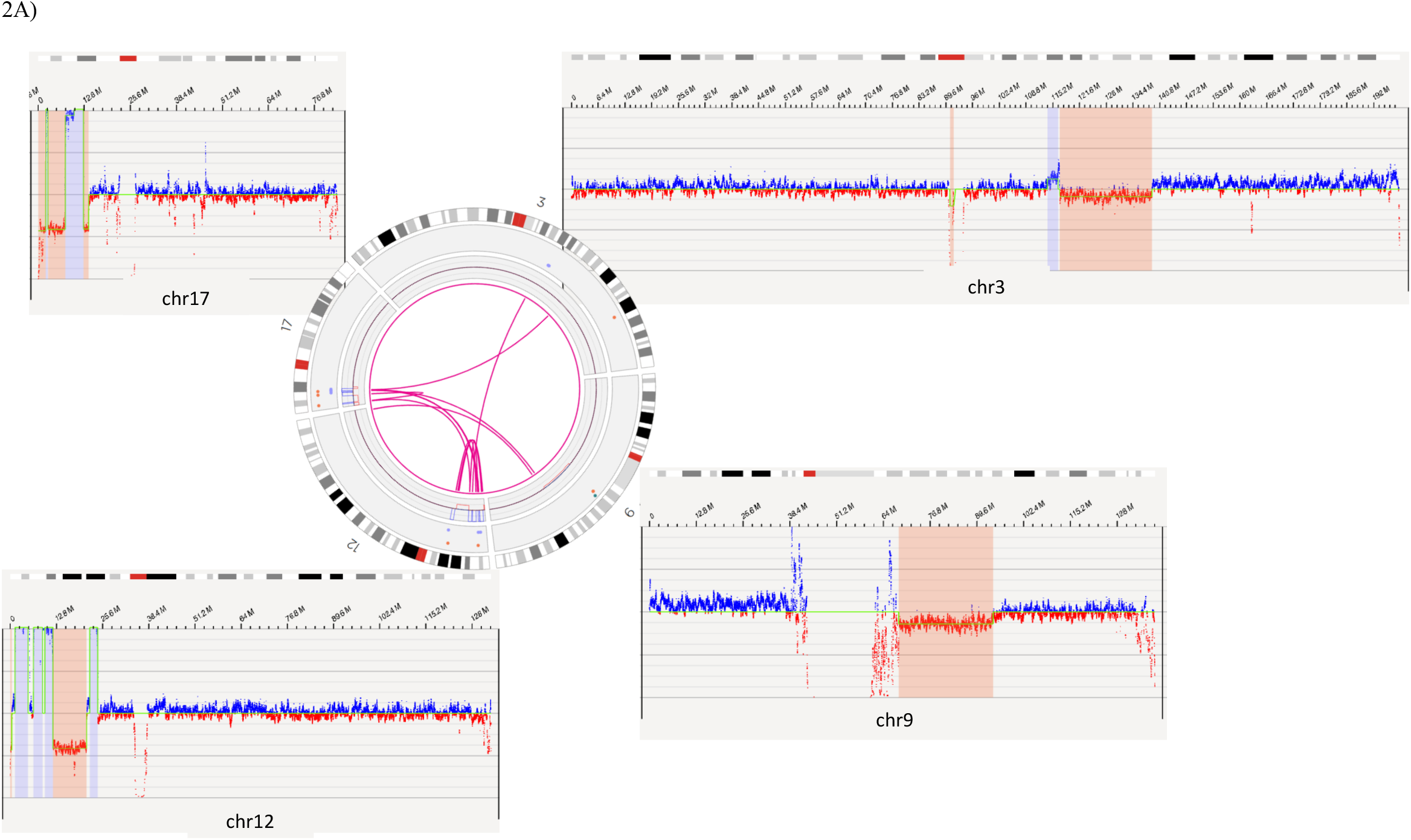
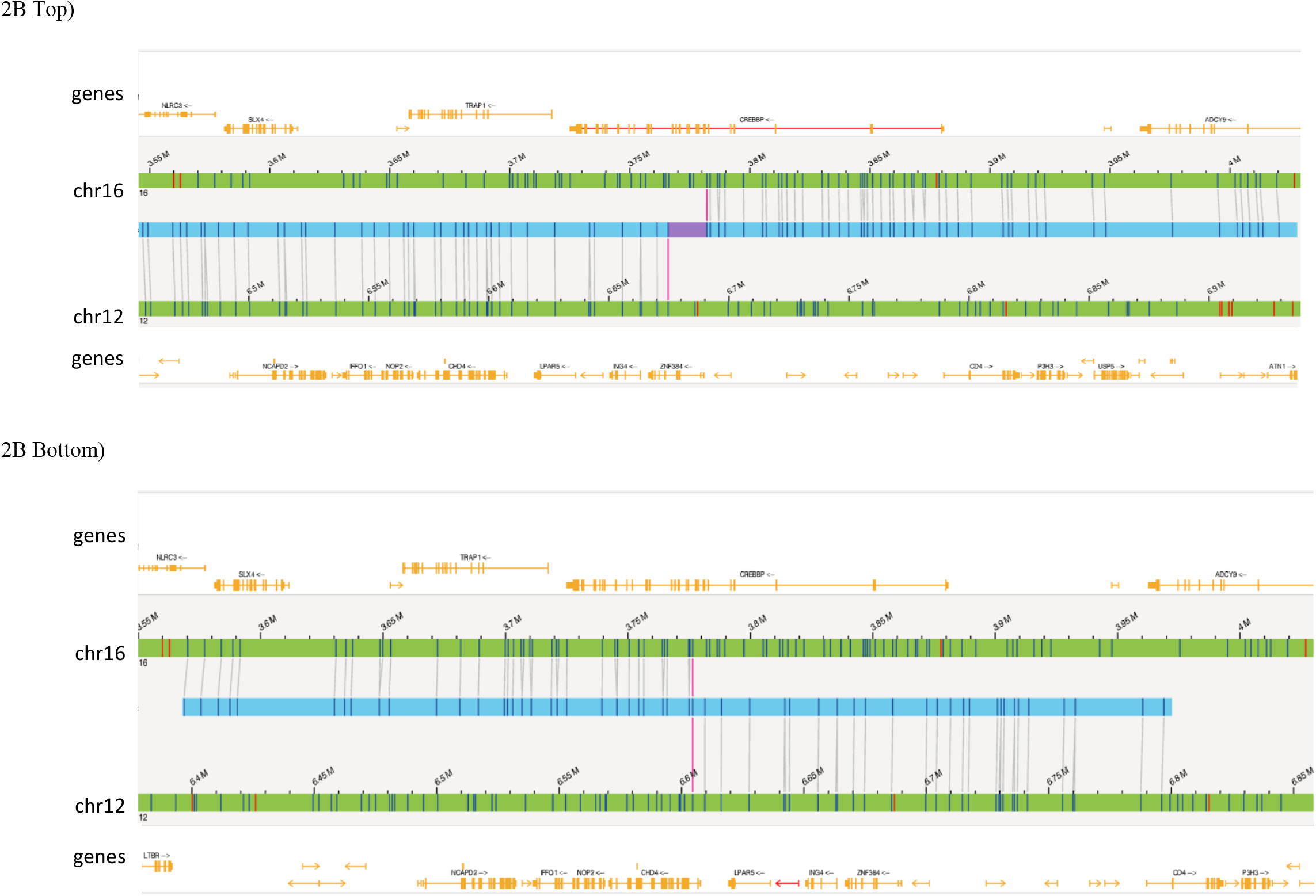
OGM captures additional novel Tier 1 or 2 findings not detected by SOC methods. 3A) Complex rearrangements that consist of a deletion of the *GATA2* gene on 3q13.31 to 3q22.3 and multiple variants on 17p13 seen in case BNGOHM-0000149. The circos plot and coverage profile of chromosomes 3, 9, 12 and 17 are shown, all of which are associated with the 17p rearrangements. 3B) The *CREBBP-ZNF384* fusion (Top) and its reciprocal translocation (Bottom) captured by OGM in case BNGOHM-0000335. The first picture shows the conjoining of chr16 with chr12 such that *CREBBP* would be fused with *ZNF384*, and second shows the reciprocal fusion point.

All incidental SVs that were detected in 18 healthy donor blood samples were compared to both a list of 206 targets/types recommended for testing by medical associations (NCCN, WHO and NHS, Supplementary Table 3) and a comprehensive list of cancer genes. Overall, tier 1 or 2 heme-associated variants were only found in two of the healthy donor samples. A loss of Y was detected in BNGOHM-0000076 and a 71 kbp deletion in 9p24.1 impacting the *PDCD1LG2* gene was identified in another donor (anonymized ID: 1000107245). Both variants were subsequently confirmed by CMA as true positive calls. In summary, based on this analysis there were no false negative SVs detected in the genomes of healthy donors, thus indicating 100% specificity (for tier 1 and 2 SVs).

Reproducibility was performed utilizing four cases, and the concordance for each SV was determined by calculating the fraction of replicates (N≥6) in which each variant was accurately identified (Supplementary Table 4). Overall, the reproducibility assay included an aggregate of 227 SVs (14 aneusomies, 46 duplications, 33 insertions, 58 deletions, 12 inversions, 32 intra-chromosomal fusions, 32 inter-chromosomal translocations). In total, there was 96% reproducibility among replicates for all SVs, CNVs and aneuploidies. A total of five variants were not detected across all replicates: two duplications, two inversions, and one insertion. The lack of detection was attributed to the SV having one of the following: in a region with segmental duplications, low variant confidence score, or high complexity in the region.

Two cell lines (KG-1 and MV4-11) were used to evaluate the limit of detection (LOD) of OGM. Two trisomies, three duplications, nine deletions, an inversion, and three translocations were assessed in the dilution series (Supplementary Table 5). In summary, at the current coverage of >1500 Gbp map-based SV calling can detect deletions at ≥ 5.0% VAF, inversions at ≥ 4.1% VAF, and translocations at ≥ 4.8% VAF. The read depth CNV calling algorithm showed LOD 10.5% VAF for gains, 15.9% VAF for losses, and 11.0% VAF for trisomies (Table 3).

**Table 3.** Limit of detection for different SV types.

| **Variant type** | **LOD** |
| --- | --- |
| Deletion | 5.0% VAF |
| Translocation | 4.8% VAF |
| Inversion | 4.1% VAF |
| CN loss | 15.9% VAF |
| CN gain | 10.5% VAF |
| Trisomy | 11.0% VAF |

## Discussion

Medical guidelines by professional societies such as the NCCN, WHO, NHS, etc recommend specific testing for SVs as part of the workup of suspected hematologic malignancies both, at diagnosis, and during follow-up testing (disease monitoring). Current laboratory practices rely on several complementary cytogenetic techniques to accomplish recommended testing, but these methods suffer from the limitation of low resolution, thereby potentially missing actionable SVs. Cytogenetic/cytogenomic testing laboratories often have customized testing algorithms depending on the disease diagnosis and stage, physician preferences and the institution’s testing and reimbursement policies. These decades long disparities typically lead to different combination of FISH and/or KT testing depending on the laboratory infrastructure, instrumentation, and availability of trained cytogenetics laboratory professionals. There is a need for innovative technologies that can overcome the challenges of traditional SOC testing and introduce a comprehensive, uniform combination of tests being performed on a routine basis. This study demonstrates that OGM can be easily implemented in the clinical setting and can substantially reduce operational complexity and improve detection rate by providing a reproducible and robust alternative to the three predominant cytogenic methods (KT, FISH, and CMA) in routine workup of most of the hematological malignancies.

A wide variety of hematologic malignancies were included during this LDT validation process and of the 148 reported SVs detected by SOC methods, OGM detected 100% of these variants. OGM was validated for different SV classes such as deletions, inversions, and translocations, all performing at or above 5% VAF threshold. Importantly, the detection of novel, actionable, clinically significant SVs solely by OGM in a significant fraction of cases (37% of study cases) adds substantial diagnostic benefits from OGM testing. One important feature of a robust assay is reproducibility and OGM detected over 96% of variants across all replicates within and between runs. It is important to note that the legacy cytogenetic methods also suffer from significant subjectivity in SV classification since KT results are highly dependent on the quality of the metaphase band resolution (usually in the 300-500 band level range for heme).

Another unique feature of the OGM assay involves the uniform pre-analytical and analytical steps, irrespective of the disease subtype, which allows for the workflow standardization and scalability for single or multiple hematologic malignancy subtypes. The incorporation of guideline-driven, disease subtype-specific target variants into the analytical process allows for a semi-automated and easy classification and reporting of variants. Additionally, if there is ambiguity in the clinical symptoms of a heme malignancy sub-type at the time of the physician consult, a “pan-heme” genomic analysis can still be conducted simultaneously in a disease-agnostic fashion.

Multiple studies, including the present study, demonstrate that OGM overcomes multiple limitations of SOC testing^11,17^. In agreement with recent published studies, this study demonstrates increased detection of clinically relevant SV by OGM compared to SOC in 37% of cases (tier 1 or tier 2 SVs that were missed by SOC). Additionally, OGM provides a standardized data acquisition and analysis process with a software solution that allows the seamless and systematic implementation and adoption across multiple laboratories.

This clinical validation is the second published study in the USA according to CAP/CLIA guidelines^18^. After the validation was completed, a representative report template was created with Tier 1A variants highlighted on page 1 and Tier 1b/2 variants listed on page 2 (Figure 3). Section A is analogous to a FISH panel with present/absent indicated for each abnormality. Section B represents a whole genome analysis and displays the SVs detected on a chromosome-by-chromosome basis (analogous to a KT).

**Figure 3.**
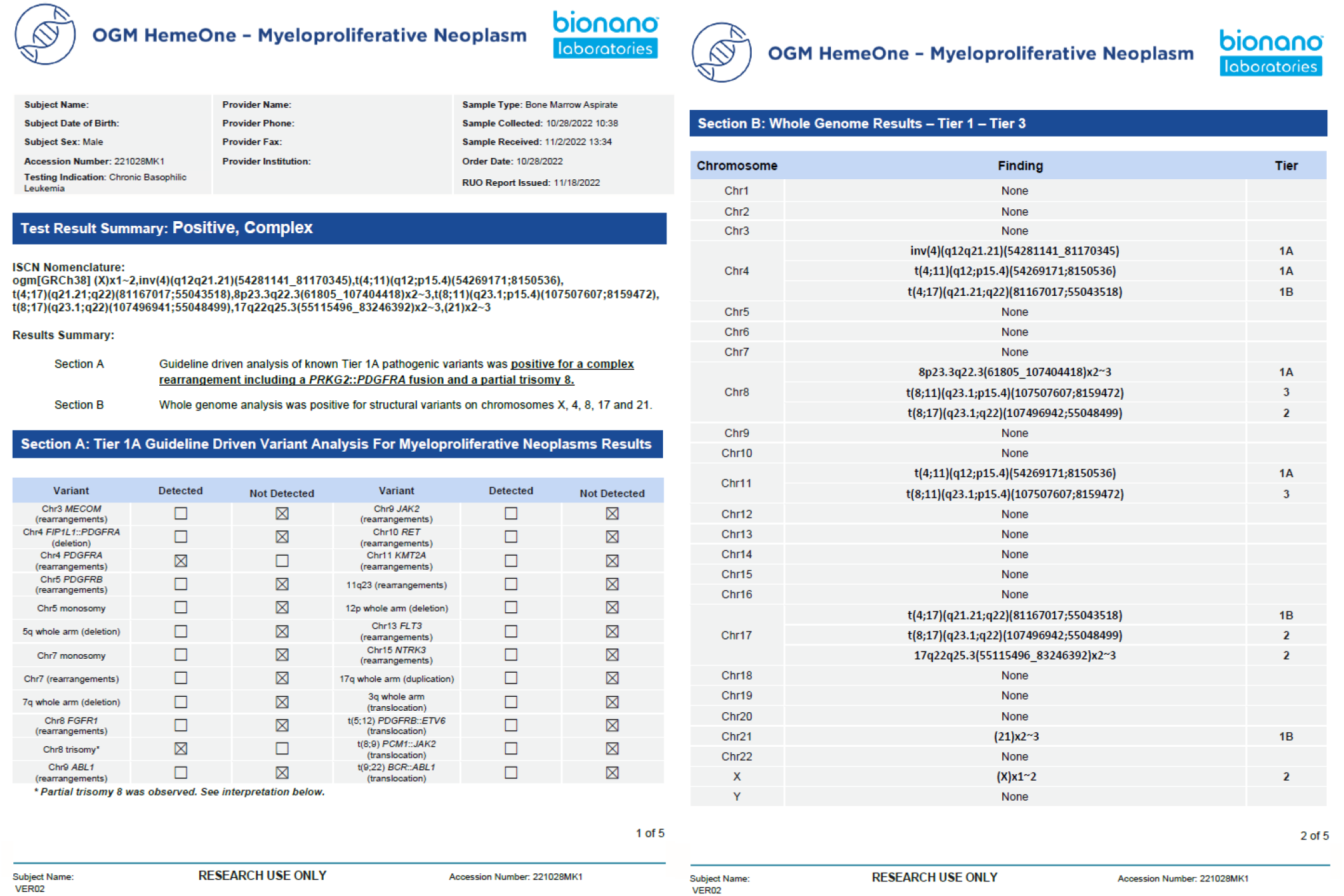
Example of a representative clinical OGM report. After demographic information, test results summary tells if any variant has been detected (positive/negative) and if the genome is simple or complex. Section A shows the defined tier 1A abnormalities as a panel and section B shows the whole genome profile from chromosomes 1-22, X and Y.

This sample-to-answer workflow established in a CLIA laboratory environment has been provided to three labs laboratories and data has been collected and analyzed according to the established standard operating procedures (data not shown). Results of the initial analysis demonstrate the potential of OGM to provide a more uniform and systematic approach to cytogenomics, which will be further evaluated in the ongoing larger multi-site clinical trial underway.

## Conclusions

OGM provides a solution that enables an alternative to multiple methods such as, KT, FISH, and CMA addressing several lacunae of the traditional methods. The OGM workflow provides an end-to-end solution from DNA isolation to the downstream SV analysis and interpretation using an analysis software to facilitate adoption in a clinical laboratory. Since the OGM assay does not require culturing of the clinical specimens, the typically observed culture biases in a cytogenetic laboratory are eliminated. OGM also allows for a sample to answer to be obtained in 4-5 days, which is extremely beneficial for the management and treatment of these patients. OGM not only detects all classes of SVs but provides the highest resolution attainable till date, by any cytogenetic method in clinical use. The ability of OGM to detect both recurrent SVs and novel fusions positions it as a first-tier test for the detection of all classes of SVs in majority of the hematological malignancies.

## Data Availability

Data will be made available upon reasonable request and in accordance with IRB protocols.

## Data Availability

Data will be made available upon reasonable request and in accordance with IRB protocols.

IRB and consent IRB-20212956 – Bionano Genomics Inc., San Diego, CA, USA

IRB-00007527 – University of Rochester Medical Center Office for Human Subject Protection

IRB-A-#00000150 (HAC IRB # 611298) - Medical College of Georgia, Augusta University, Augusta, GA, USA

## Disclosures

AWCP, KK, AS, JH, BC, JE, AC, TS, AH, AC are all salaried employees of Bionano Genomics

RK has received honoraria, and/or travel funding, and/or research support from Illumina, Asuragen, QIAGEN, Perkin Elmer Inc, Bionano Genomics, Agena, Agendia, PGDx, Thermo Fisher Scientific, Cepheid, and BMS.

## Acknowledgements

Acknowledgements: The authors wish to acknowledge Bionano clinical team: Alvin Yee, Mike Gallagher, James Yu, Vruti Mehta, Kelsea Chang, Shuk Shukor, Alma Lastrella, Beth Matthews, Sean Guy, Anusha Mylavarapu, Julia Brushett, Evelyn Crescini, Sheng-Wei Chang for their support with data management, training, conducting validation experiments and analysis.

## Funding

This study was funded in part by Bionano Genomics, inc.

## Supplementary Figures

**Supplementary Figure 1.**
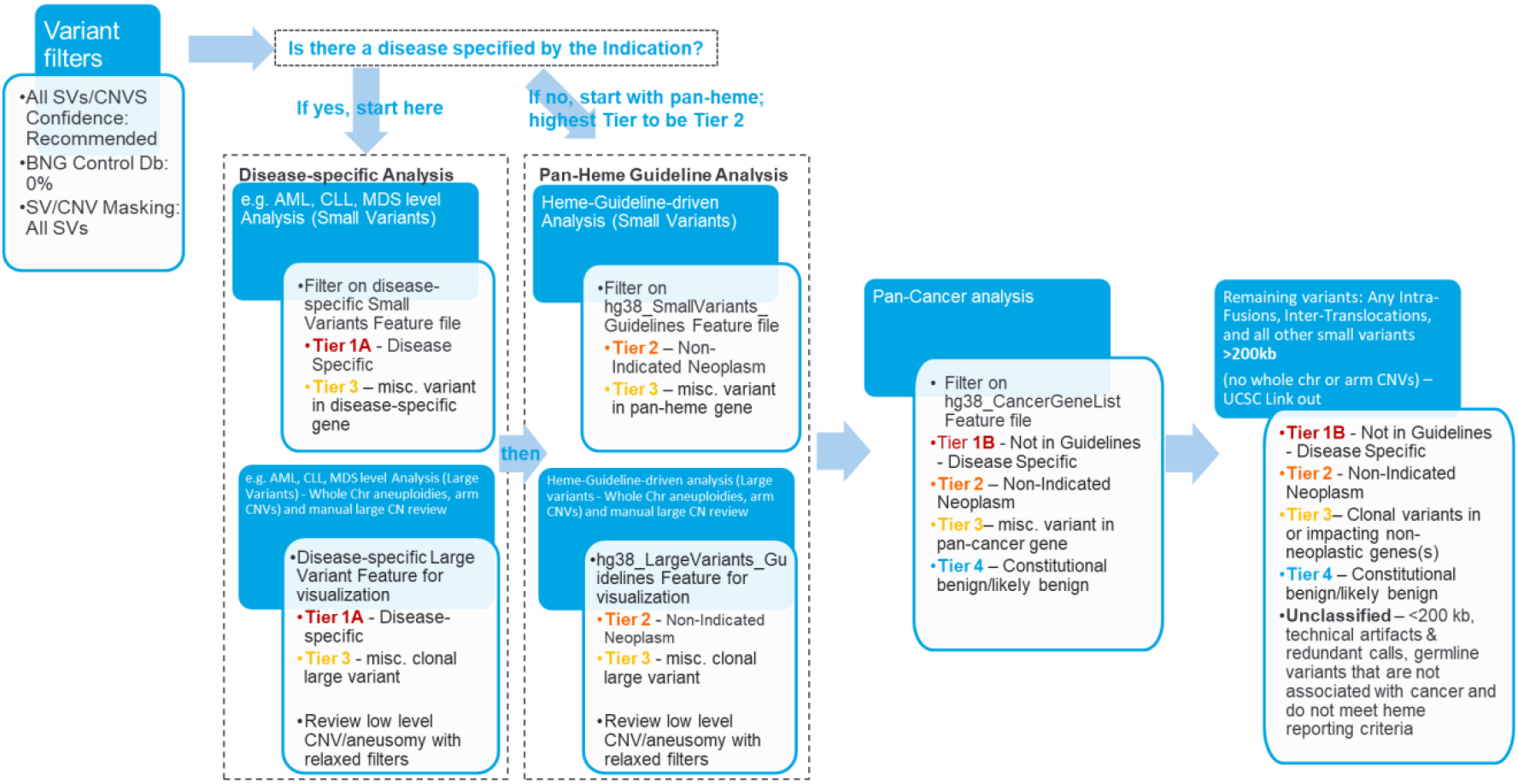
Overview of analysis and interpretation workflow. The curation and classification process consists of four steps: 1) disease-specific: Several organizations such as the World Health Organization (WHO), the National Health Service (NHS) and the National Comprehensive Cancer Network (NCCN) have published guidelines for assessing clinically relevant genomic regions for hematological malignancies. The analysts apply a filter to select for variants overlapping known loci associated with the disease of the sample (e.g. AML), and then classify the selected SVs as Tier 1A or 3; 2) Pan-hematological cancers: The analyst removes the first filter, and applies another filter to select for SVs seen in hematological cancers and classifies them as Tier 2 or 3; 3) Pan-cancer: SVs overlapping other cancer-associated genes are selected and classified as Tier 1B, 2, 3 or 4; and 4) Remaining variants: All remaining SVs that were fusions, or were >200 kbp are classified as Tier 1B, 2, 3 or 4.

**Supplementary Table 1.** Summary of molecule quality statistics for 207 data points. *Overall, 185 out of 207 data points passed the recommended molecule quality metrics.

| Analytical Performance Metrics (N = 207 data points) * |  |  |  |  |  |  |
| --- | --- | --- | --- | --- | --- | --- |
| Molecule N50 |  |  | Map rate |  | Effective coverage |  |
| Average N50 (kbp) | >230 kbp | >200 kbp | Average Map Rate (%) | >70% | Average Effective Coverage (x) | > 320x |
| 261.15 kbp | 89% | 100% | 87.16% | 99% | 434.61x | 98% |

Supplementary Table 2. OGM detects additional novel Tier 1 or 2 findings not seen in SOC platforms. This table contains a list of the disease-associated findings of karyotype, FISH, chromosomal microarray, and OGM. Novel OGM findings have been confirmed by orthogonal methods. The OGM circos plots show an overview of somatic SVs identified in the tumor genomes. Also, karyotyping variants annotated as “add” cannot be compared for concordance because of insufficient information. *Any OGM ISCN entries with an appended asterisk indicate that a microarray was run for confirmation, and those OGM variants with asterisks were confirmed.

See file: Supplemental_table_2.xlsx

**Supplementary Table 3.** List of unique NCCN, WHO and NHS structural variants across hematological malignancies that were assessed as part of the review process.

| Chr | target | Rearr. | Del. | Dup. | Transl_intrachr | Transl_inter | Inversion | Trisomy | Monosomy | CNV |
| --- | --- | --- | --- | --- | --- | --- | --- | --- | --- | --- |
| 1 | *ABL2* | X |  |  |  |  |  |  |  |  |
| 1 | *BCL10* | X |  |  |  |  |  |  |  |  |
| 1 | *CDKN2C* |  | X |  |  |  |  |  |  |  |
| 1 | *CKS1B* |  |  | X |  |  |  |  |  |  |
| 1 | *JAK1* | X |  |  |  |  |  |  |  |  |
| 1 | *LMNA::NTRK1* |  |  |  | X |  |  |  |  |  |
| 1 | *NTRK1* | X |  |  |  |  |  |  |  |  |
| 1 | *TAL1* | X | X |  |  |  |  |  |  |  |
| 1 | *TPR::NTRK1* |  |  |  | X |  |  |  |  |  |
| 1 | whole arm, p |  | X |  |  |  |  |  |  |  |
| 1 | whole arm, q |  |  | X |  |  |  |  |  |  |
| 1 | 1q21 |  |  | X |  |  |  |  |  |  |
| 1;2 | *TPM3::ALK* |  |  |  |  | X |  |  |  |  |
| 1;7 | *RNF11::BRAF* |  |  |  |  | X |  |  |  |  |
| 1;7 | *TRB::TAL1* |  |  |  |  | X |  |  |  |  |
| 1;7 | *MIGA1::BRAF* |  |  |  |  | X |  |  |  |  |
| 1;8 | *ETV3::NCOA2* |  |  |  |  | X |  |  |  |  |
| 1;14 | *BCL10::IGH* |  |  |  |  | X |  |  |  |  |
| 1;19 | *PBX1::TCF3* |  |  |  |  | X |  |  |  |  |
| 1;22 | *RBM15::MRTFA* |  |  |  |  | X |  |  |  |  |
| 2 | *ALK* | X |  |  |  |  |  |  |  |  |
| 2 | *EML4::ALK* |  |  |  | X |  |  |  |  |  |
| 2 | *IGK* | X |  |  |  |  |  |  |  |  |
| 2 | *REL* | X |  |  |  |  |  |  |  |  |
| 2;5 | *NPM1::ALK* |  |  |  |  | X |  |  |  |  |
| 2;8 | *IGK::MYC* |  |  |  |  | X |  |  |  |  |
| 2;10 | *KIF5B::ALK* |  |  |  |  | X |  |  |  |  |
| 2;17 | *CLTC::ALK* |  |  |  |  | X |  |  |  |  |
| 3 | 3q26.2 | X |  |  |  |  |  |  |  |  |
| 3 | *BCL6* | X |  |  |  |  |  |  |  |  |
| 3 | *FOXP1* | X |  |  |  |  |  |  |  |  |
| 3 | *GATA2::MECOM* |  |  |  | X |  |  |  |  |  |
| 3 | *MECOM* | X |  |  |  |  | X |  |  |  |
| 3 | *TP63* | X |  |  |  |  |  |  |  |  |
| 3 | whole chrom |  |  |  |  |  |  | X |  |  |
| 3 | whole arm, q | X |  |  | X |  |  |  |  |  |
| 3;5 | *NPM1::MLF1* |  |  |  |  | X |  |  |  |  |
| 3;8 | *MYC::BCL6* |  |  |  |  | X |  |  |  |  |
| 3;14 | *FOXP1::IGH* |  |  |  |  | X |  |  |  |  |
| 4 | *FIP1L1::PDGFRA* |  | X |  | X |  |  |  |  |  |
| 4 | *PDGFRA* | X |  |  |  |  |  |  |  |  |
| 4 | whole chrom |  |  |  |  |  |  | X |  |  |
| 4;11 | *KMT2A::AFF1* |  |  |  |  | X |  |  |  |  |
| 4;14 | *FGFR3::IGH* |  |  |  |  | X |  |  |  |  |
| 4;14 | *IGH* |  |  |  |  | X |  |  |  |  |
| 4;14 | *NSD2::IGH* |  |  |  |  | X |  |  |  |  |
| 5 | *CSF1R* | X |  |  |  |  |  |  |  |  |
| 5 | *EBF1* |  | X | X |  |  |  |  |  |  |
| 5 | *PDGFRB* | X |  |  |  |  |  |  |  |  |
| 5 | *TLX3* | X |  |  |  |  |  |  |  |  |
| 5 | whole chrom |  |  |  |  |  |  | X | X |  |
| 5 | whole arm, q |  | X |  |  |  |  |  |  |  |
| 5 | 5q31.2 |  | X |  |  |  |  |  |  |  |
| 5;11 | *NUP98::NSD1* |  |  |  |  | X |  |  |  |  |
| 5;12 | *PDGFRB::ETV6* |  |  |  |  | X |  |  |  |  |
| 5;14 | *BCL11B::TLX3* |  |  |  |  | X |  |  |  |  |
| 6 | *CCND3* | X |  |  |  |  |  |  |  |  |
| 6 | *DUSP22::IRF4* |  |  |  | X |  |  |  |  |  |
| 6 | *DUSP22* | X |  |  |  |  |  |  |  |  |
| 6 | *IRF4* | X |  |  |  |  |  |  |  |  |
| 6;7 | p25.3;q32.3 |  |  |  |  | X |  |  |  |  |
| 6;9 | *DEK::NUP21* |  |  |  |  | X |  |  |  |  |
| 6;11 | *KMT2A::AFDN* |  |  |  |  | X |  |  |  |  |
| 6;14 | *IGH::CCND3* |  |  |  |  | X |  |  |  |  |
| 7 | *BRAF* | X |  |  |  |  |  |  |  |  |
| 7 | *EZH2* |  | X | X |  |  |  |  |  |  |
| 7 | *IKZF1* | X | X | X |  |  |  |  |  |  |
| 7 | *TRB* | X |  |  |  |  |  |  |  |  |
| 7 | *TRG* | X |  |  |  |  |  |  |  |  |
| 7 | whole chrom | X |  |  |  |  |  |  | X |  |
| 7 | whole arm, q |  | X | X |  |  |  |  |  |  |
| 7 | 7q31.2 |  | X |  |  |  |  |  |  |  |
| 7;10 | *TRB::TLX1* |  |  |  |  | X |  |  |  |  |
| 7;11 | *NUP98::HOXA13* |  |  |  |  | X |  |  |  |  |
| 7;12 | *MNX1::ETV6* |  |  |  |  | X |  |  |  |  |
| 7;19 | *CLIP3::BRAF* |  |  |  |  | X |  |  |  |  |
| 7;22 | *PACSIN2::BRAF* |  |  |  |  | X |  |  |  |  |
| 8 | *FGFR1* | X |  |  |  |  |  |  |  |  |
| 8 | *LYN* | X |  |  |  |  |  |  |  |  |
| 8 | *MYC* | X |  |  |  |  |  |  |  |  |
| 8 | whole chrom | X |  |  |  |  |  | X |  |  |
| 8;9 | *PCMI::JAK2* |  |  |  |  | X |  |  |  |  |
| 8;14 | *IGH::MYC* |  |  |  |  | X |  |  |  |  |
| 8;18 | *MYC::BCL2* |  |  |  |  | X |  |  |  |  |
| 8;21 | *RUNX1::RUNX1T1* |  |  |  |  | X |  |  |  |  |
| 8;22 | *IGL::MYC* |  |  |  |  | X |  |  |  |  |
| 9 | *ABL_extraPh* |  |  | X |  |  |  |  |  |  |
| 9 | *ABL1* | X |  | X |  |  |  |  |  |  |
| 9 | *CD274* | X | X | X |  |  |  |  |  |  |
| 9 | *CDKN2A* |  | X | X |  |  |  |  |  |  |
| 9 | *CDKN2B* |  | X | X |  |  |  |  |  |  |
| 9 | *JAK2* | X |  |  |  |  |  |  |  |  |
| 9 | *NUP214::ABL1* |  |  |  | X |  |  |  |  |  |
| 9 | *PAX5* |  | X | X |  |  |  |  |  |  |
| 9 | *PDCD1LG2* | X | X | X |  |  |  |  |  |  |
| 9 | whole chrom |  |  |  |  |  |  | X |  |  |
| 9 | whole arm, q |  | X |  |  |  |  |  |  |  |
| 9;11 | *KMT2A::MLLT3* |  |  |  |  | X |  |  |  |  |
| 9;22 | *BCR::ABL1* |  |  |  |  | X |  |  |  |  |
| 10 | *RET* | X |  |  |  |  |  |  |  |  |
| 10 | *TLX1* | X |  |  |  |  |  |  |  |  |
| 10 | whole chrom |  |  |  |  |  |  | X |  |  |
| 10;11 | *KMT2A::MLLT10* |  |  |  |  | X |  |  |  |  |
| 10;14 | *TLX1::TRD* |  |  |  |  | X |  |  |  |  |
| 11 | whole arm, q |  | X | X |  |  |  |  |  |  |
| 11 | *ATM* |  | X |  |  |  |  |  |  |  |
| 11 | *CCND1* | X |  |  |  |  |  |  |  |  |
| 11 | *KMT2A* | X |  |  |  |  |  |  |  |  |
| 11 | *LMO2* | X |  |  |  |  |  |  |  |  |
| 11 | *NUP98* | X |  |  |  |  |  |  |  |  |
| 11 | 11q23 | X |  |  |  |  |  |  |  |  |
| 11;14 | *11q13::IGH* |  |  |  |  | X |  |  |  |  |
| 11;14 | *CCND1::IGH* |  |  |  |  | X |  |  |  |  |
| 11;14 | *TRD::LMO1* |  |  |  |  | X |  |  |  |  |
| 11;18 | *BIRC3::MALT1* |  |  |  |  | X |  |  |  |  |
| 11;19 | *KMT2A::ELL* |  |  |  |  | X |  |  |  |  |
| 11;19 | *KMT2A::MLLT1* |  |  |  |  | X |  |  |  |  |
| 12 | *BTG1* |  | X | X |  |  |  |  |  |  |
| 12 | *CCND2* | X |  |  |  |  |  |  |  |  |
| 12 | *ETV6* |  | X | X |  |  |  |  |  |  |
| 12 | whole chrom |  |  |  |  |  |  | X | X |  |
| 12 | whole arm, p |  | X |  | X |  |  |  |  |  |
| 12;21 | *ETV6::RUNX1* |  |  |  |  | X |  |  |  |  |
| 13 | *DLEU2* |  | X |  |  |  |  |  |  |  |
| 13 | *DLEU7* |  | X |  |  |  |  |  |  |  |
| 13 | *FLT3* | X |  | X |  |  |  |  |  |  |
| 13 | *RB1* |  | X | X |  |  |  |  |  |  |
| 13 | whole chrom |  |  |  |  |  |  |  | X |  |
| 13 | 13q14 |  | X |  |  |  |  |  |  |  |
| 14 | *IGH* | X |  |  |  |  |  |  |  |  |
| 14 | *TCL1A/B* | X |  |  |  |  |  |  |  |  |
| 14 | *TRA* | X |  |  |  |  |  |  |  |  |
| 14 | *TRA_TRD* | X |  |  |  |  |  |  |  |  |
| 14 | *TRD* | X |  |  |  |  |  |  |  |  |
| 14 | whole chrom |  |  |  |  |  |  |  | X |  |
| 14 | 14q32 |  | X |  |  |  |  |  |  |  |
| 14;16 | *16q23::IGH* |  |  |  |  | X |  |  |  |  |
| 14;16 | *IGH::MAF* |  |  |  |  | X |  |  |  |  |
| 14;18 | *IGH::BCL2* |  |  |  |  | X |  |  |  |  |
| 14;18 | *IGH::MALT1* |  |  |  |  | X |  |  |  |  |
| 14;20 | *IGH::MAFB* |  |  |  |  | X |  |  |  |  |
| 14;20 | *IGH* |  |  |  |  | X |  |  |  |  |
| 14;20 | *MAFB::IGH* |  |  |  |  | X |  |  |  |  |
| 15 | *NTRK3* | X |  |  |  |  |  |  |  |  |
| 15 | whole chrom |  |  |  |  |  |  | X |  |  |
| 15;17 | *PML::RARA* |  |  |  |  | X |  |  |  |  |
| 16 | *CBFA2T3::GLIS2* |  |  |  | X |  |  |  |  |  |
| 16 | *CBFB::MYH11* |  |  |  | X |  |  |  |  |  |
| 17 | *TP53* |  | X |  |  |  |  |  |  |  |
| 17 | whole chrom |  |  |  |  |  |  | X | X |  |
| 17 | whole arm, p |  | X |  | X |  |  |  |  |  |
| 17 | whole arm, q |  |  | X |  |  |  |  |  |  |
| 17 | iso17q_wholearm |  |  | X |  |  |  |  |  |  |
| 17;19 | *TCF3::HLF* |  |  |  |  | X |  |  |  |  |
| 18 | *BCL2* | X |  |  |  |  |  |  |  |  |
| 18 | *MALT1* | X |  |  |  |  |  |  |  |  |
| 18 | whole chrom |  |  |  |  |  |  | X |  |  |
| 19 | *EPOR* | X |  |  |  |  |  |  |  |  |
| 19 | *JAK3* | X |  |  |  |  |  |  |  |  |
| 19 | *TYK2* | X |  |  |  |  |  |  |  |  |
| 19 | whole chrom |  |  |  |  |  |  | X |  |  |
| 20 | whole arm, q |  | X |  |  |  |  |  |  |  |
| 21 | *RUNX1_amp* |  |  | X |  |  |  |  |  |  |
| 21 | *RUNX1* |  |  |  |  |  |  |  |  | X |
| 22 | *BCR_extraPh* |  |  | X |  |  |  |  |  |  |
| 22 | *IGL* | X |  |  |  |  |  |  |  |  |
| 22 | whole chrom |  |  |  |  |  |  | X |  |  |
| X | idic(X)_parm |  |  | X |  |  |  |  |  |  |
| X | idic(X)_qarm |  | X |  |  |  |  |  |  |  |
| X | *CFLR2* | X | X | X |  |  |  |  |  |  |
| X | *CSF2RA* |  | X | X |  |  |  |  |  |  |
| X | *IL3RA* |  | X | X |  |  |  |  |  |  |
| Y | whole chrom |  | X |  |  |  |  |  |  |  |

**Supplementary Table 4.** Reproducibility by SV type.

| Sample | SV Type | Genomic region | Size (kbp) | Concordance |
| --- | --- | --- | --- | --- |
| BNGOHM-0000178 | CN Loss | 4q22.2q28.1 | 35868 | 6/6 |
|  | Deletion | 1q21.1 | 18 | 6/6 |
|  | Deletion | 23q23 | 36 | 6/6 |
|  | Duplication | 4q22.1 | 230 | 6/6 |
|  | Insertion | 2p25.2 | 41 | 6/6* |
|  | Interchr Transl. | t(7;12)(q21.12;p11.21) | -- | 6/6 |
|  | Interchr Transl. | t(13;17)(q14.3;p13.2) | -- | 6/6 |
|  | Intrachr Transl. | t(7;7)(q11.21;q32.2) | -- | 6/6 |
|  | Intrachr Transl. | t(20;20)(q11.21;q13.33) | -- | 6/6 |
| BNGOHM-0000181 | CN Gain | 14q12q21.3 | 15162 | 6/6 |
|  | CN Loss | 3p14.2 | 736 | 6/6 |
|  | Deletion | 4p14 | 17 | 6/6 |
|  | Duplication | 6p25.1 | 568 | 6/6 |
|  | Insertion | 8q22.3 | 7 | 5/6 |
|  | Inversion | 6q16.1 | 134 | 5/6 |
|  | Inversion | 3q26.1 | 102 | 6/6 |
|  | Interchr Transl. | t(6;10)(q12;q24.2) | -- | 6/6 |
|  | Intrachr Transl. | t(6)(q12;q16.1) | -- | 6/6 |
| BNGOHM-0000185 | CN Loss | 16q23.2q23.3 | 2840 | 7/7 |
|  | Deletion | 17q24.2 | 130 | 7/7 |
|  | Duplication | 10q11.22 | 707 | 2/7 |
|  | Duplication | 22q11.22 | 278 | 6/7 |
|  | Insertion | 12q22 | 16 | 6/7 |
|  | Interchr Transl. | t(1;22)(q21.2;q11.22) | -- | 7/7 |
| BNGOHM-0000195 | CN Gain | 4q25q35.2 | 79440 | 7/7 |
|  | Aneuploidy/Gain | 8 | 120787 | 7/7 |
|  | Aneuploidy/Gain | 11 | 126084 | 7/7 |
|  | CN Loss | 16q22.1 | 2046 | 7/7 |
|  | Deletion | 14q13.2 | 23 | 7/7 |
|  | Duplication | 9p23 | 449 | 7/7 |
|  | Insertion | 4q24 | 152 | 7/7 |
|  | Insertion | 7q11.21 | 56 | 7/7 |
|  | Interchr Transl. | t(2;4)(q37.3;q25) | -- | 7/7 |
|  | Intrachr Transl. | t(5;5)(q21.3;q33.1) | -- | 7/7 |
|  | Intrachr Transl. | t(7;7)(p12.1;q31.1) | -- | 7/7 |

Supplementary Table 5. Details of Limit of detection for different variant SV types.

See file: Supplemental_table_5.xlsx

## Notes

### Author Declarations

IRB and consent IRB 20212956 Bionano Genomics Inc., San Diego, CA, USA IRB 00007527 University of Rochester Medical Center Office for Human Subject Protection IRB A 00000150 (HAC IRB 611298) Medical College of Georgia, Augusta University, Augusta, GA, USA

